# Sequential data assimilation of the stochastic SEIR epidemic model for regional COVID-19 dynamics

**DOI:** 10.1101/2020.04.13.20063768

**Authors:** Ralf Engbert, Maximilian M. Rabe, Reinhold Kliegl, Sebastian Reich

## Abstract

Newly emerging pandemics like COVID-19 call for predictive models to implement precisely tuned responses to limit their deep impact on society. Standard epidemic models provide a theoretically well-founded dynamical description of disease incidence. For COVID-19 with infectiousness peaking before and at symptom onset, the SEIR model explains the hidden build-up of exposed individuals which creates challenges for containment strategies. However, spatial heterogeneity raises questions about the adequacy of modeling epidemic outbreaks on the level of a whole country. Here, we show that by applying sequential data assimilation to the stochastic SEIR epidemic model, we can capture the dynamic behavior of outbreaks on a regional level. Regional modeling, with relatively low numbers of infected and demographic noise, accounts for both spatial heterogeneity and stochasticity. Based on adapted models, short-term predictions can be achieved. Thus, with the help of these sequential data assimilation methods, more realistic epidemic models are within reach.

---

The initial spread of the novel coronavirus in Germany [1] resulted in containment measures based on reduced traveling and social distancing [3]. In epidemic standard models [2, 14], which provide a dynamical description of epidemic outbreaks [7, 22], containment measures aim at a reduction of the contact parameter. Since the contact parameter is one of the critical parameters that determine the speed of increase of the number of infectious individuals, estimating the contact parameter is a key basis for epidemic modeling [18].

From early on, the situation of COVID-19 has been characterized by extreme spatial heterogeneity [1]. In the initial phase of the outbreak, spatial heterogeneity was caused by random travel-based imports of infectious cases and enhanced by local events with increased contacts; after introduction of non-pharmaceutical interventions, spatial heterogeneity was sustained. Therefore, over the full observation period, the assumption of homogeneous mixing must be relaxed [12], and coupled dynamics of regional models seem to be a more adequate description [16]. However, when modeling a relatively small region with a population size of *N* = 10^5^, compared to the country level with populations of *N* = 10^7^ to 10^9^, one must address the problem of stochasticity [9, 12] (see The stochastic SEIR model). The combination of dynamical modeling and substantial fluctuations calls for sequential data assimilation methods for parameter inference [15, 20] as widely used, for example, in numerical weather prediction [5].

We investigate how the stochastic SEIR epidemic model [2] applies to regional data of COVID-19 incidence under non-pharmaceutical interventions, i.e., where epidemic dynamics were confined to regions and coupling between them could be neglected. The model assumes *S, E, I*, and *R* compartments representing susceptible, exposed, infectious, and recovered individuals (Fig. 1). This model is particularly important for the description of the spread of COVID-19, since infectiousness seems to peak on or before symptom onset [13] and models without the exposed compartment cannot adequately address the time delay between the build-up of exposed and infectious individuals.

**Figure 1:**
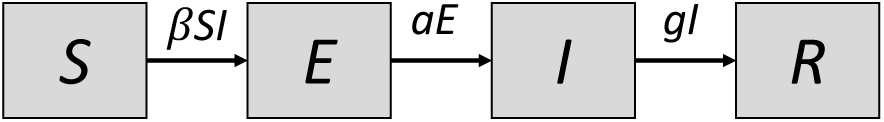
The SEIR model. The population is divided into four compartments that represent susceptible, exposed, infectious, and recovered individuals. The contact parameter *β* is critical for disease transmission, 1/*a* and 1/*g* are the average durations of exposed and infectious periods, resp. Unlike in the standard model, the birth and death processes are neglected in short-term simulations discussed throughout the current study.

Since we are interested in short-term modeling (weeks to months), we neglect birth and death processes as a first-order approximation for the dynamics of the model. The disease-related model parameters are the rate parameters *a* = 1/*Z* (with an average latency period *Z*) and *g* = 1/*D* (with a mean infectious period *D*), which can be estimated independently from the analysis of infected cases [13, 16]. Therefore, the time-dependent contact parameter *β* is the most critical parameter that needs to be determined via data assimilation [20]. It is directly related to the basic reproductive number *R*_0_ in a SEIR-type model (see SEIR model and basic reproductive number). Therefore, non-pharmaceutical interventions that aim at *R*_0_ < 1 translate into the relation *β* < *g* in the model.

In the following, we will use a combination of sequential data assimilation and stochastic modeling on the regional level to estimate the spatial heterogeneity in the spread of epidemics and to show how to use such a combined approach for epidemics predictions and uncertainty quantifications.

## Results

The key motivation of the current study was to apply sequential data assimilation to the stochastic SEIR model to estimate the contact parameter. We successfully applied an ensemble Kalman filter [10, 15, 20] to recover the contact parameter from simulated data (see Parameter recovery from simulated data). When applied to empirical data on a regional level, the estimation of the contact parameter produces a comparable evidence profile (see Application to empirical data).

In the early phase of the COVID-19 outbreak in Germany, the reported cumulative numbers of cases increased rapidly (Fig. 2a,b), however, the epidemic dynamics vary from region to region. This spatial heterogeneity is due to the different onset times of the disease in different regions, but it is also enhanced by variations in the local contact parameters *β*. In response to the containment measures, we expect *β* to change over time.

**Figure 2:**
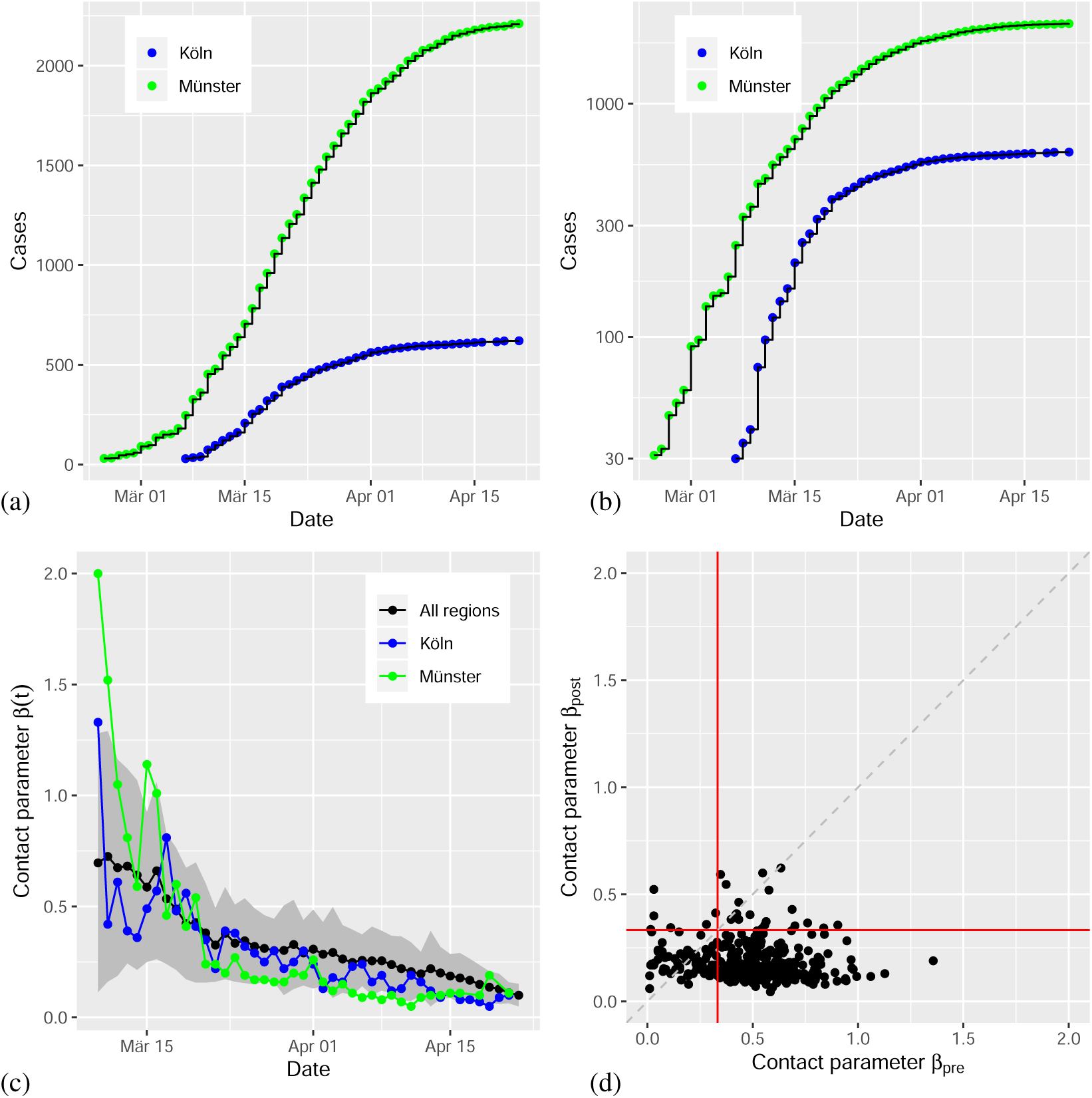
Analysis of best fit time-dependent contact parameters *β*_∗_(*t*_*k*_); date refers to report of case at RKI. (a) For two regions (LK Köln and LK Münster), the cumulative numbers show a strong increase after different disease onset times. (b) Semi-logarithmic scaling suggests approximate exponential growth in early as well as later regimes. (c) The time dependent contact parameter *β*_∗_(*t*_*k*_) indicates a small decrease over time due to social distancing interventions (black: average for 320 regions; red, blue: contact parameter for the examples above; grey shading: standard deviation across regions. (d) Scatter plot of the time averaged contact parameter *β*_pre_ before intervention and *β*_post_ after intervention. Note that the critical value for disease containment is *β*_crit_ = 1/3 per day in our model (red lines).

### Time-dependence of the contact parameter

An estimation of the time-dependence of the contact parameter is achieved via the model’s best fit. An approximative instantaneous negative log-likelihood *L*(*t*_*k*_, *β*) of the contact parameter *β* at observation time *t*_*k*_ is obtained from the ensemble Kalman filter (see Model inference based on sequential data assimilation). Thus, by determining the minimum of *L*(*t*_*k*_, *β*) with respect to *β* at time *t*_*k*_ we estimate the time-dependence of the best fit *β\**(*t*_*k*_) (Fig. 2c). The black line represents the average time dependence for all 320 regions included in the analysis; standard deviations are indicated by the grey area. The results for the two example regions are given in their corresponding colors.

The non-pharmaceutical interventions to counter the spread of COVID-19 were implemented at slightly different points in time across Germany. In the majority of the regions, closings of schools and other educational institutions started on March 16, while large-scale contact bans were implemented on March 22. As a result of these social distancing measures will have an impact on the contact parameter, we expected to observe a related drop in the contact parameter over time. Before we present a corresponding analysis, it should be made clear that none of these measures can produce an immediate effect on the observed cases of infected individuals because of the latency period. Since sequential data assimilation will need several data points to adapt the model to the data, the related interval should be as long as possible to achieve a reliable estimate of the contact parameter. Therefore, we selected the average value of *β\**(*t*_*k*_) over the three days from March 17th to March 19th as a pre-intervention value. The average over March 31st to April 2nd is taken as an estimate of the post-intervention value. To analyze the effect across regions, we computed average values *β*_pre_ (March 17-19) and *β*_post_ (March 31-April 2) of the relevant *β\**(*t*_*k*_) for all regions. The resulting scatter plot indicates a clear reduction of the numerical value of the contact parameter from *β*_pre_ to *β*_post_ (Fig. 2d). The reduction is statistically significant (Wilcoxon test, *p* < 0.01).

### Simulations with time-dependent contact parameter

The contact parameter *β* is the most critical parameter determining the dynamics of the stochastic SEIR model. After a time-resolved estimation of the best fit *β*_∗_(*t*_*k*_), we are able to generate simulations from an initial state to predict the future trajectory (Fig. 3). Simulations I begin with the first epidemic day in the corresponding region with greater than or equal to 30 cases. The initial number of infected *I*_0_ is set to the observed number of cases *y*_obs_(*t*_0_), while the initial number of exposed is set to *E*_0_ = *g*/*a* · *I*_0_, which would hold at epidemic equilibrium for *m* > 0 (for *m* = 0 both *E* and *I* tend to zero with *E*/*I* ≈ *g*/*a*). The initial number of infected was disturbed by noise representing uncertainties in the initial model states. Forward iterations with the estimated time-varying contact parameter show that the slope of the epidemic curve is approximately reproduced by the model (Fig. 3a,c; grey lines indicate the ensemble of simulated trajectories; blue points are observed data).

**Figure 3:**
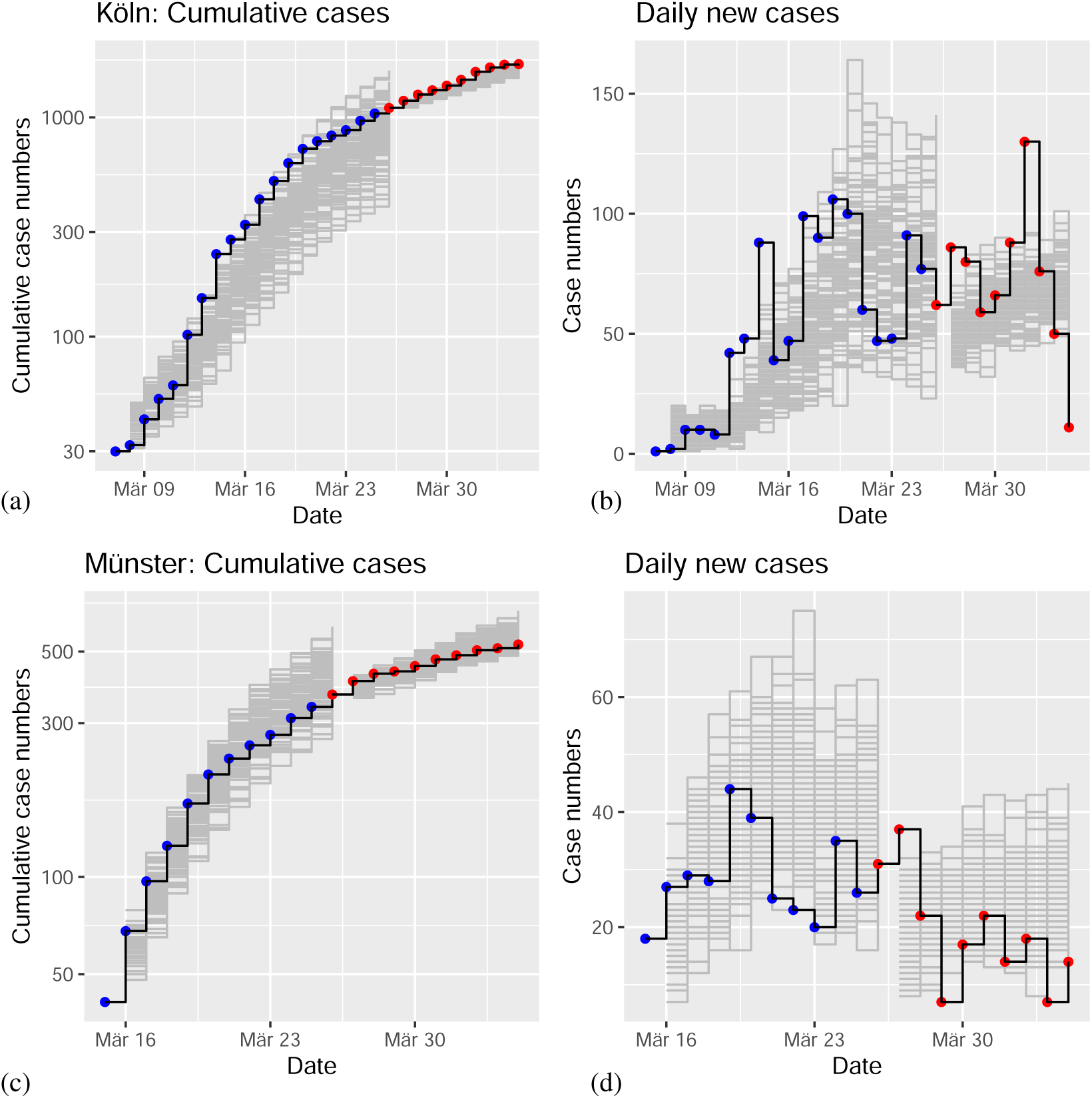
Simulations of the stochastic SEIR model for two example regions. Simulations I indicate an ensemble of 100 runs of the model with initial conditions from the first epidemic day with number of cases greater than or equal to 30 (grey: ensemble of trajectories; blue: observations). Simulations II start at March 26, using an ensemble size of 100 after data assimilation (grey: ensemble of trajectories; red: observations). (a) Cumulative cases of infected individuals over time for LK Köln. Daily reported new cases for Köln. (c) Cumulative cases for LK Münster. (d) Daily new cases for Münster. Date refers to report of case at RKI.

Simulation II starts at March 26 and exploits the full potential of sequential data assimilation. The sequential data assimilation approach via the ensemble Kalman filter (see Ensemble Kalman filter) is based on the forward modeling of an ensemble of trajectories. After each time step (1 day), the ensemble of trajectories is compared to the next observation and adjusted via a linear regression step. Thus, we obtained an adapted ensemble of internal model states for each epidemic day. Here we exploit this fact to run a forward simulation with initial conditions from the assimilated ensemble of internal model states. The corresponding forward simulations are close to the real time-evolution of the epidemics in the two example regions (Fig. 3a,c; grey lines indicate the ensemble of simulated trajectories; blue points are observed data). A related plot of the daily reported new cases indicates an approximately constant level of numbers of new cases for Köln (Fig. 3b) and slowly decreasing daily new cases for Münster (Fig. 3d); both predictions are in agreement with empirical observations.

### Predictions for two different scenarios

The forward simulations discussed in the previous section demonstrated the predictive power of the SEIR model when using sequential data assimilation. In the next step, we generated simulations under two different scenarios. In scenario I, we started with the adapted ensemble of internal model states after data assimilation (April 16) and iterated the model forward with the mean contact parameter estimated from the period of April 14 to April 16, that is well after interventions were implemented (Fig. 4, green area). The simulations continue to match the time-course of infected cases for both example regions (Fig. 4a,b). Daily reported case numbers show a decline for both regions (Fig. 4c,d).

**Figure 4:**
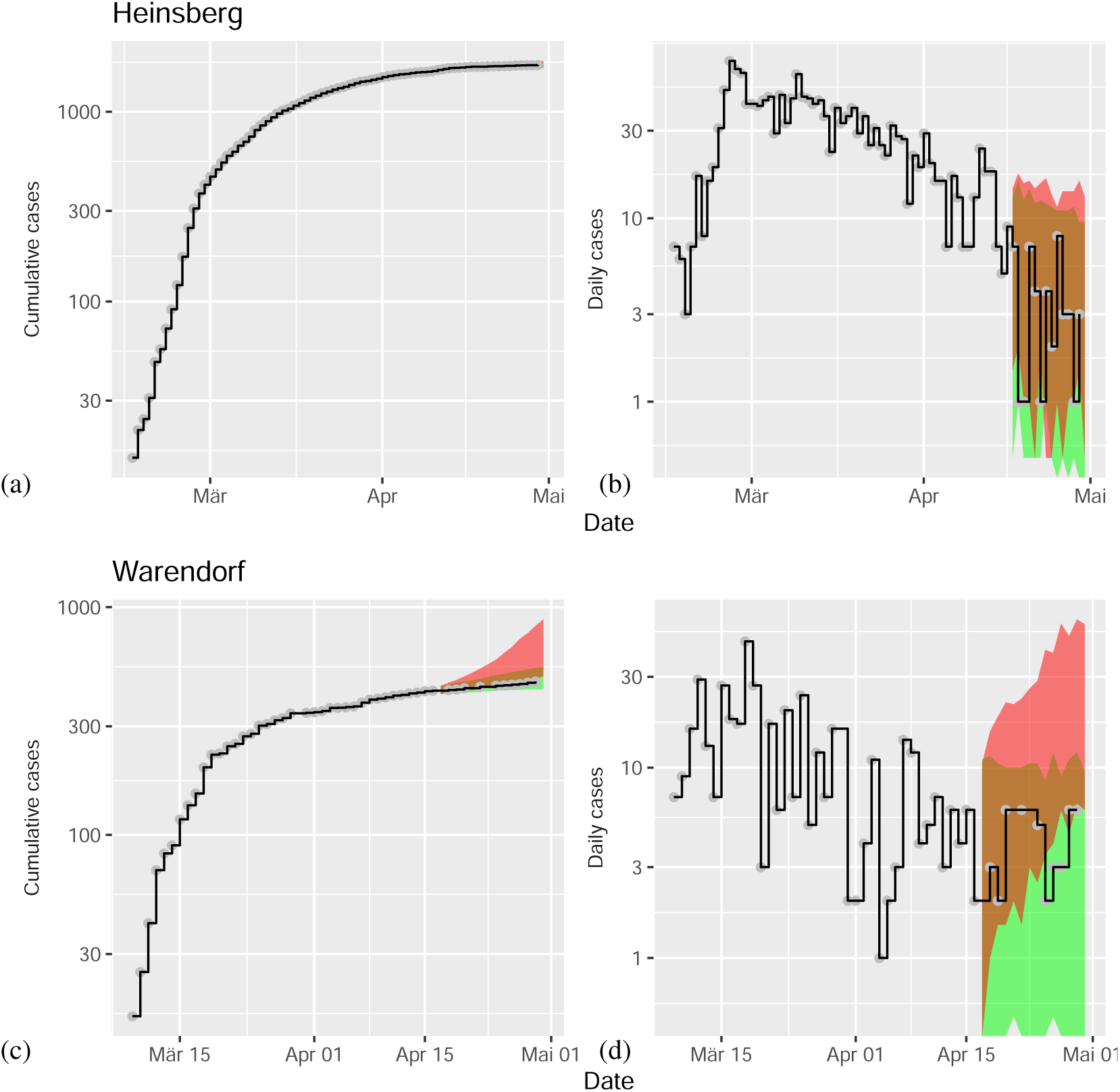
Model predictions for COVID-19 after data assimilation in comparison to data (black lines). In scenario I (green area), an assimilated ensemble of internal model states starts the forecast with contact parameter *β*_post_ (continuation of social distancing interventions). In scenario II (red area), the equivalent forecast is generated with contact parameter *β*_pre_ (termination of interventions). Predictions for cumulative case numbers in Heinsberg. (b) Predictions of daily new cases in Heinsberg. (c) Predictions of cumulative cases for Warendorf. (d) Predictions of daily new cases for Warendorf. Date refers to report of case at RKI.

In scenario II, we assumed that all governmental intervention measures had been terminated. Therefore, we used the estimated contact parameters from the period of March 17 to March 19. Again, we started simulations with the adapted ensemble of internal states after sequential data assimilation (Fig. 4, red area). For both example regions, we observe a strong increase in infected cases under scenario II (Fig. 4c,d). This dramatic increase can be seen most clearly in the plot of daily numbers of new cases (for more examples see Supplementary Information Appendix).

## Discussion

The ongoing worldwide spread of the new coronavirus exerts enormous pressure on healthcare systems, societies and governments. Therefore, predicting the epidemic dynamics under the influence of non-pharmaceutical interventions (NPI) is an important problem from a data science and mathematical modeling perspective [19]. The motivation of the current work was to explore the potential of sequential data assimilation [15, 20] to create a regional epidemic model as a forecasting tool.

The standard epidemic SEIR-type models implement a compartmental description under the assumption of homogeneous mixing of individuals [2]. More realistic modeling approaches must account for spatial heterogeneity due to time-varying disease onset times, regionally different contact rates, and the time-dependence of the contact rates due to the implementation of containment strategies. However, these regional descriptions require models that include the effects of demographic stochasticity due to the limited size of populations and the low number of cases in the region considered [6]. The effects of such statistical fluctuations are inherently reproduced via stochastic versions of the standard epidemic models [9, 12].

We have demonstrated the potential of sequential data assimilation to reproduce COVID-19 dynamics at the level of a regional, stochastic model. With the help of the ensemble Kalman filter [10], we successfully recovered the contact parameter from the simulated data and obtained reliable estimates from the empirical data. The contact parameter is the most critical free parameter in the stochastic SEIR model, since the other parameters (mean exposed and infectious duration) can be estimated independently from observed time series [13, 16]. Moreover, the contact parameter of the SEIR model is directly related to the basic reproductive rate *R*_0_ [17]. Therefore, our approach could also be framed as a model-based method for statistical inference of the basic reproductive number.

Next, we ran a time-resolved data assimilation that generated estimates of the time-dependence of the contact parameter. The drop in mean contact rates from an early (*β*_pre_, March 17 to 19) to a later period (*β*_post_, March 31 to April 2) indicates the effect of non-pharmaceutical interventions. We also generated model prediction for two different scenarios. In scenario I, simulations produce forecasts with start date April 16. The previously assimilated ensemble provide the initial conditions and the contact parameter is set to the value estimated for the post-intervention period from April 14 to April 16. In scenario II, we replaced the post-intervention contact parameter with its pre-intervention value, estimated from the data for the period March 17 to March 19. As a result, the two scenarios predict rather different temporal developments (decline of daily new cases for scenario I and strong increase for scenario II). Therefore, our model predictions suggest that lifting of the non-pharmaceutical interventions could potentially turn the epidemic dynamics back to the exponential increase from before their implementation. Such predictions can easily be scaled up to the federal state level (Bundesländer) or to the country level; a corresponding predictive model will be potentially quite robust due to its explicit modeling of spatial and temporal heterogeneities, captured by a separate time-course of the contact parameter for each region.

A recent simulation study by Li et al. [16] used a similar approach of sequential data assimilation for dynamic epidemic models. However, they implemented a deterministic SEIR model and extended it with additional noise assumptions. We proposed the usage of the stochastic SEIR model in the formulation of a master equation [9] which can be simulated exactly and numerically efficiently using Gillespie’s algorithm [11]. A more complex spatiotemporal stochastic model has been considered in [4].

Furthermore, the state-parameter estimation in [16] utilises the ensemble Kalman filter directly on an augmented state space [20]. Contrary to that study, we found a direct application of the ensemble Kalman filter to the augmented state space (*X, β*) not suitable because of the strongly nonlinear interaction between the model states *X* and the contact parameter *β*. This led us to consider a two stage approach which combines the ensemble Kalman filter for state estimation with a likelihood based inference of the contact parameter *β* [20].

Our current study was mainly motivated by the methodological problem of a possible contribution from data assimilation to epidemics modeling based on a stochastic SEIR model. There are obvious limitations within our current modeling framework, which we did not address because of our methodological focus. Longer-term predictions (∼ months) are important, but they critically depend on an estimation of undocumented infections (see Li et al. [16]). Such hidden infections create, after recovery, an unknown reduction in the number of susceptibles, which slows down the epidemic dynamics; such an effect is currently not included in our current model. However, it seems compatible with our framework to extend the SEIR model by an additional class of undocumented infected individuals [16].

Another important limitation of these results comes from the simplifying assumption that there is no coupling to neighboring regions. As a consequence, the regional differences in the contact parameter could be at least partly due to differences in the contacts between the regions. Couplings between the regions [16] could also be integrated into our modeling framework. However, the non-coupling approximation might be realistic in the situation of social distancing and travel bans during the period investigated here.

## Methods

### RKI data on COVID-19 in Germany

The Robert Koch Institute (RKI), the central scientific institution in the field of biomedicine within the portfolio of the Federal Ministry of Health, provides daily access to the number of confirmed cases, deaths, and recovered patients, broken down into 412 counties, six age groups, and by gender. As they are official records, only cases certified by doctors or labs according to a strict medical protocol in accordance with the Infection Protection Act are entered into the data base. The exact time of an infection is usually not known. The associated time stamp refers to the date on which the local health authority became aware of the case and recorded it electronically. As records are passed from the physician or lab via local and state health authorities to the RKI, there is a delay of several days before cases are reported on the website. Thus, the statistics relating to the most recent three or four days are incomplete and cannot be interpreted; retrospective updates and corrections are made for all days of the pandemic spell as they become available. Also the date at which the case is reported at the RKI (i.e., the Erkrankungsdatum/date of illness) is not more recent than the date at which the infection occurred. We use data up to and including April 30, as reported on June 22, 2020; they are included as part of the supplement.

### SEIR model and basic reproductive number

The SEIR epidemic model is a four-compartment model with susceptibles (individuals who are able to contract the disease), exposed (individuals who are infected but not yet infectious), infectious, and recovered (individuals who are immune). The model is typically formulated as a system of ordinary differential equations (ODE), i.e.,

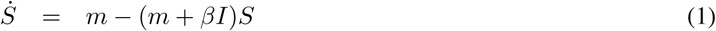

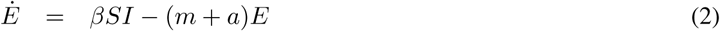

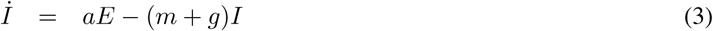

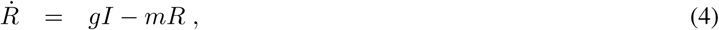

where the total number of individuals *N* = *S* + *E* + *I* + *R* is constant under temporal evolution due to 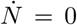. The ODE system, Eq. (1-4), has a non-trivial equilibrium point, denoted as epidemic equilibrium (*S*_0_, *E*_0_, *I*_0_, *R*_0_), where the number of susceptibles *S*_0_ at equilibrium is related to the basic reproductive number [8]. Since we aim at a short-term description of the system, we neglect birth and death processes here, equivalent to the limit *m* → 0, we obtain

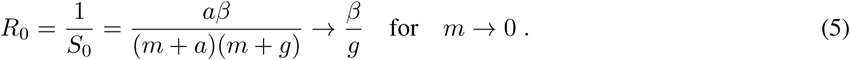

We use a numerical values of *g* = 1/3 per day, equivalent to an average infectious period of *D* = 3 days [16]. As a consequence, the critical condition for disease containment *R*_0_ < 1 is obtained for *β* < *β*_crit_ = 1/3 per day in our model. The median latency period has been estimated as 5.2 days [13]. Here we used a numerical value of *a* = 1/5.2 per day for the rate parameter of the exposed individuals.

### The stochastic SEIR model

While the classical model is formulated as a system of ordinary differential equations, we are exploring its application to a relatively low number of cases in the early phase of the current epidemics on the regional level with population sizes from 10^5^ to 10^6^. Therefore, we use the stochastic SEIR model in the form of a master equation [9], which is particularly useful for modeling small numbers of infected individuals occuring in smaller regions or at the beginning of an epidemic.

The demographic SEIR model contains four variables denoted by *X* = (*S, E, I, R*)^T^ ∈ ℕ^4^, representing the number of individuals in each of the four classes with a constant population size *N* = *S* + *E* + *I* + *R*. The transition rate of the ODE compartmental model translates into transition probabilities in the master equation formulation for the evolution of the model’s conditional probability density, that is,

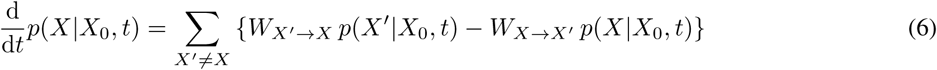

with transition probabilities given in Table 1 and initial condition *X*_0_. The individual trajectories for the model’s temporal evolution can be simulated exactly and numerically efficiently [9] using Gillespie’s algorithm [11].

**Table 1:** Transitions and transition probabilities in the stochastic SEIR model. The transition are from state *X* = (*S, E, I, R*)^T^ to *X*^*t*^ with probability *W*_*X→X*_*t*.

| $X'$ | | | | $W_{X \rightarrow X'}$ |
| --- | --- | --- | --- | --- |
| $S - 1$ | $E + 1$ | $I$ | $R$ | $\beta SI/N$ |
| $S$ | $E - 1$ | $I + 1$ | $R$ | $aE$ |
| $S$ | $E$ | $I - 1$ | $R + 1$ | $gI$ |

### Model inference based on sequential data assimilation

Publicly available data on the cumulative number of infected individuals is used to infer the model states *X* = (*S, E, I, R*)^T^ and the contact parameter *β* of the stochastic SEIR model. Note that the cumulative number of infected individuals corresponds to *Y* = *I* + *R* in the SEIR model.

In the present study, we combine sequential data assimilation for the model states with an approximate log-likelihood function for the contact parameter [20]. The basic algorithmic idea is to propagate an ensemble of *M* model forecasts using Gillespie’s algorithm up to the next available observation point *t*_*k*_. The forecast ensemble is denoted by 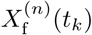 with *n* ∈ {1, …, *M*}. We used an ensemble size of *M* = 100 in this study. The reported cumulative number of infected individuals *y* (*t*) is then used via a linear regression approach to obtain the adjusted model states 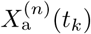. This step is implemented via the ensemble Kalman filter in its formulation of [21, 20]. While the forecast ensemble is used to compute the temporary negative log-likelihood *L*(*t*_*k*_, *β*) of the model’s contact parameter *β* at time *t*_*k*_, the adjusted model states serve as starting values for the next Gillespie prediction cycle.

The above algorithm is run with a fixed range of contact parameters *β* ∈ [*β*_min_, *β*_max_] and for a fixed time window [*t*_initial_, *t*_final_] of available data points *y*_obs_(*t*_*k*_). The best fit contact parameter *β*_∗_(*t*_*k*_) at any time any *t*_*k*_ is the one that minimises the temporary negative log-likelihood function, that is,

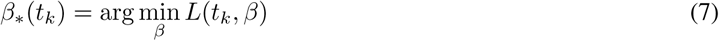

with *L*(*t*_*k*_, *β*) defined by (13) below.

### Ensemble Kalman filter

The reported cumulative number of infected individuals *y*_obs_(*t*_*k*_) is linked to the model states *X* = (*S, E, I, R*)^T^ via

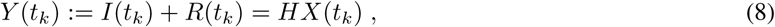

i.e., *H* = (0, 0, 1, 1). As an initial condition, we set *I*(*t*_0_) equal to the reported number of infected cases and *R*(*t*_0_) = 0 so that *y*_obs_(*t*_0_) = *I*(*t*_0_)+*R*(*t*_0_), and *E*(*t*_0_) = *g*/*a·I*(*t*_0_) with additive noise. We assume that the errors in the observed *y*_obs_(*t*_*k*_) is additive Gaussian with mean zero and variance *ρ*. We set *ρ* = 10 in our experiments. The analysis step of the ensemble Kalman filter is now based on the empirical mean

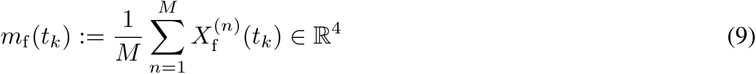

and the empirical covariance matrix

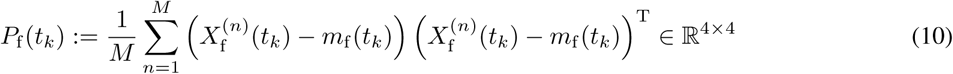

of the forecast ensemble. These two quantities are used to quantify the forecast uncertainty. Combining the forecast uncertainty with the assumed data error model leads to the linear regression formula [21, 20]

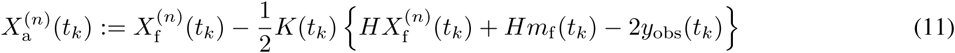

with the Kalman gain defined by

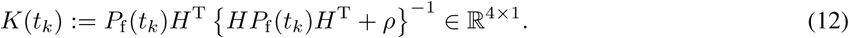

The resulting analysis ensemble 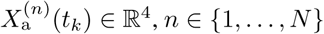, can be mapped back onto the integers ℕ^4^ if needed.

### Model evidence

The model’s negative log-likelihood at an observation time *t*_*k*_ is approximated by

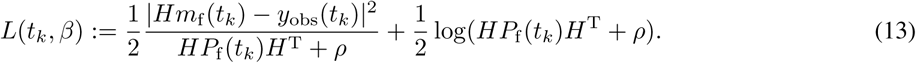

Note that the first contribution penalises the data misfit while the second penalises model uncertainty. The smaller the negative log-likelihood, the better the chosen model parameter *β* fits the data *y*_obs_(*t*_*k*_) at time *t*_*k*_. The best parameter fit over a time window [*t*_min_, *t*_max_] is defined as the value of *β* which minimises the cumulative negative log-likelihood

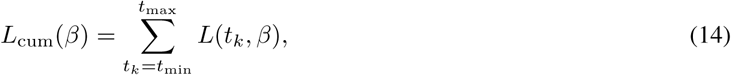

that is,

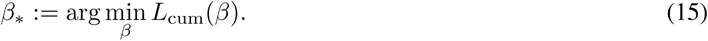

### Parameter recovery from simulated data

To test the inference scheme, we simulated data for 20 days. In Figure 5a, the black line indicates the evolution of the SEIR model’s predicted cumulative numbers of infected individuals, *Y* (*t*_*k*_) = *HX*(*t*_*k*_) = *I*(*t*_*k*_) + *R*(*t*_*k*_). As in the real data, red dots represent the daily number of reported cases. In the simulation, the contact rate was chosen as *β*_true_ = 0.6. In the following, we analyzed whether this true value could be recovered using the inference procedures described above.

**Figure 5:**
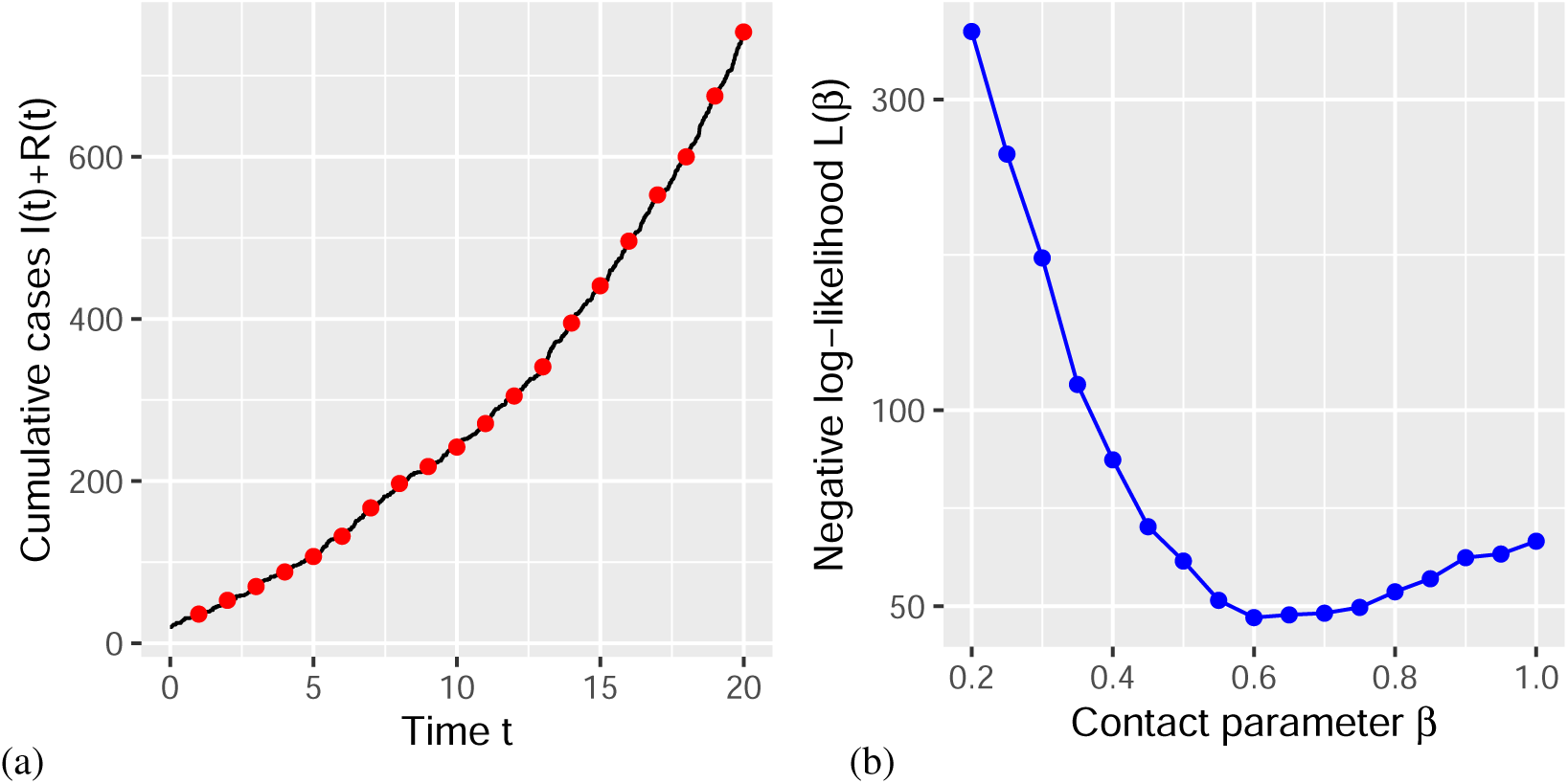
Parameter recovery analysis. (a) Simulated data with *b* = 0.6. (b) Negative log-likelihood *L*_cum_(*β*) indicates a minimum at about the true parameter value.

We varied the contact rate *β* and determined the cumulative negative log likelihood values *L*_cum_(*β*), Eq. (14). The position of the minimum of *L*_cum_(*β*) indicates the best estimate for the numerical value of the underlying contact rate *β*_∗_, Eq. (15). The position of the minimum turns out to be close to the true value, *β*_∗_ ≈ *β*_true_ = 0.6 (Fig. 5b). Thus, parameter recovery can be demonstrated for a relatively short time series of 10 observations, which represents a typical data-set in the early phase of newly emerging epidemics. Next, we apply our inference scheme to real data.

### Application to empirical data

Since the parameter inference was successful with simulated data, the next step was an application to empirical observations. We applied the inference framework to two regional data sets from the RKI data base. As an example, we selected the COVID-19 time series for Köln (RKI data, population size *N* = 1, 085, 664), which includes 27 days of observations with more than 30 cases and is plotted in Figure 6a. The parameter estimation yields an estimate for the contact rate of *β*_∗_ ≈ 0.7 (Fig. 6b). Thus, an analysis of the negative log-likelihood function produced qualitatively similar results for the simulated SEIR time series and the empirical data for a representative region. In the main text, we carry out an estimation of the time-resolved instantaneous optimal parameter values *β*_∗_(*t*_*k*_), Eq. (7), using the instantaneous negative log-likelihood function *L*(*t*_*k*_, *β*), Eq. (13).

**Figure 6:**
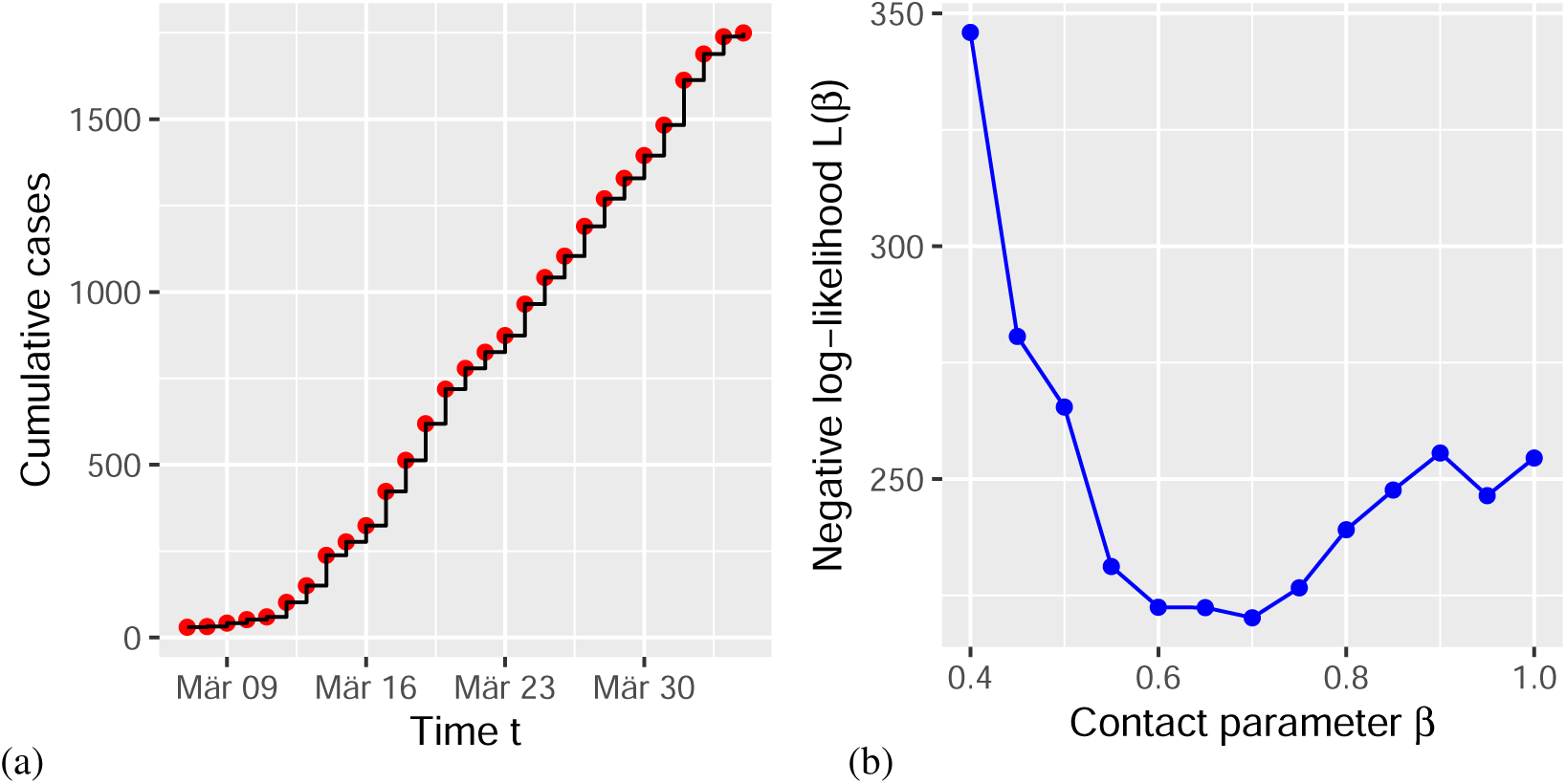
Contact parameter estimates for real data. (a) Data for Köln; date refers to report of case at RKI. (b) Negative log-likelihood *L*_total_(*β*) for Köln give a minimum at *β*_∗_ ≈ 0.7.

We found that our results were relatively insensitive to the choice of the measurement error variance *ρ* appearing in (12) and (13). At the same time, we emphasise that the errors in the reported cumulative numbers of infected individuals are complex, may vary over time, and will certainly impact on the inferred parameters. The same applies to the unknown initial model states *X*(*t*_0_) = (*S*(*t*_0_), *E*(*t*_0_), *I*(*t*_0_), *R*(*t*_0_))^T^ and their uncertainties.

## Data Availability

Data are publicly available via Robert Koch Institute (RKI), Berlin, Germany, at https://npgeo-corona-npgeo-de.hub.arcgis.com/, accessed: 2020-04-08; source code (R Language of Statistical Computing) and data used in our study are available via Open Science Framework (OSF).

https://osf.io/7dshm/

## Acknowledgments

We thank Klaus Dietz, Tübingen, for comments on the manuscript. This work was supported by a grant from Deutsche Forschungsgemeinschaft, Germany (SFB 1294, project no. 318763901).

## Data and Code Availability

Data and source code for simulations, analyses, and figures is available via Open Science Framework (OSF) at https://osf.io/7dshm/

## Supplementary Information Appendix

This PDF file includes additional examples for regional modeling.

**Figure S1:**
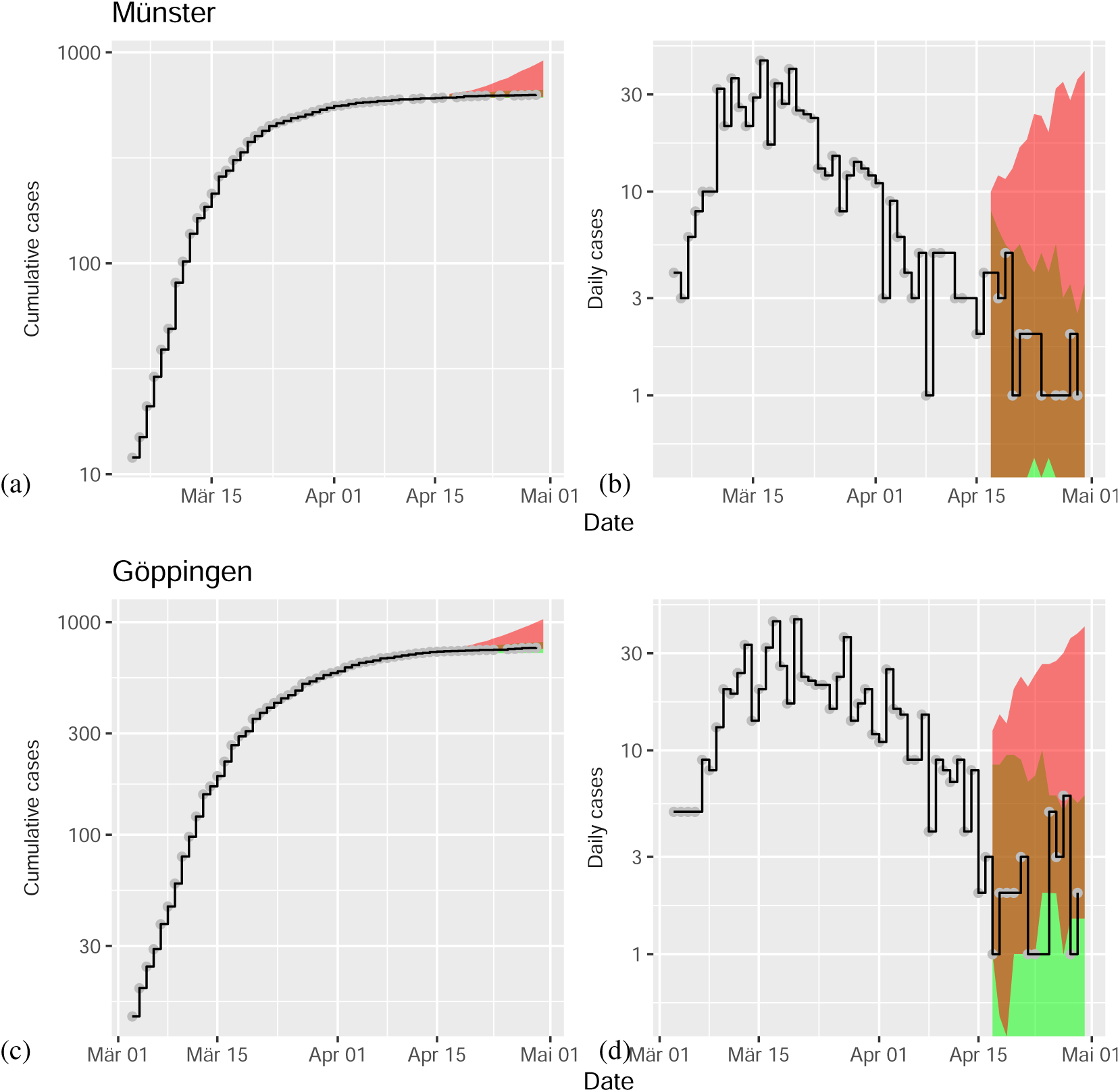
Model predictions for COVID-19 after data assimilation. For details of modeling scenarios I and II see main text and Figure 4.

**Figure S2:**
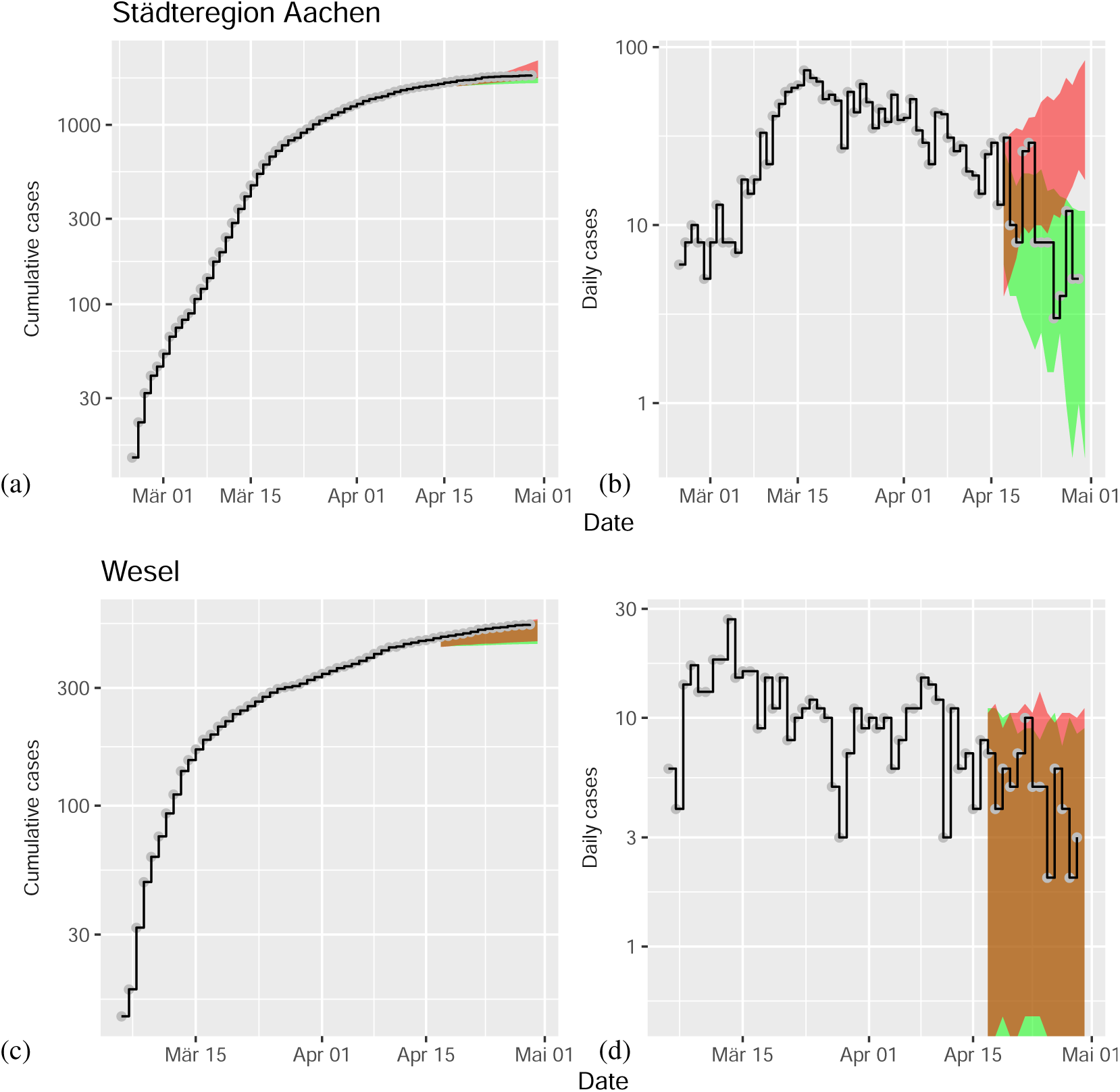
Model predictions for COVID-19 after data assimilation. For details of modeling scenarios I and II see main text and Figure 4.

**Figure S3:**
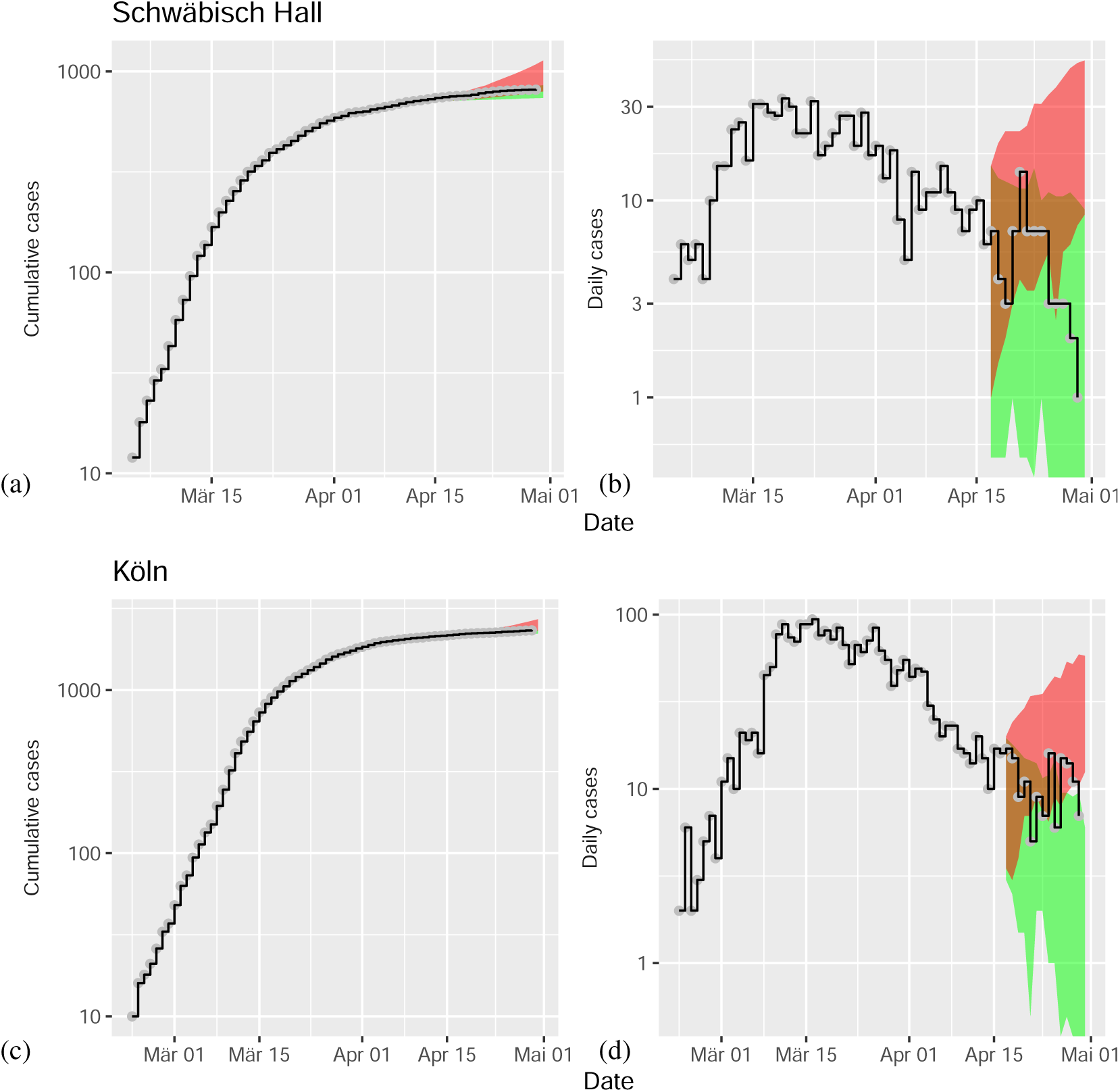
Model predictions for COVID-19 after data assimilation. For details of modeling scenarios I and II see main text and Figure 4.

**Figure S4:**
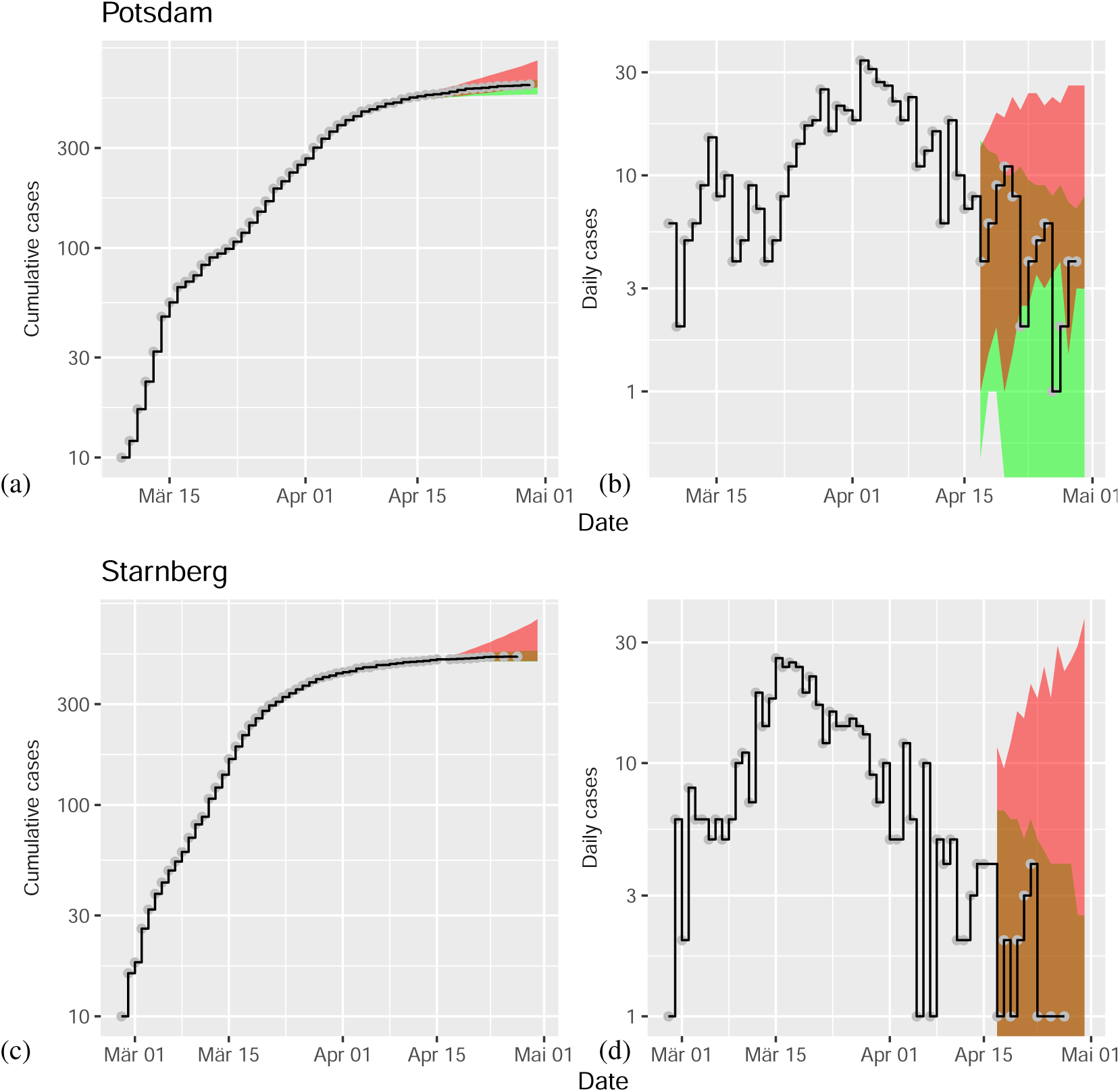
Model predictions for COVID-19 after data assimilation. For details of modeling scenarios I and II see main text and Figure 4.

**Figure S5:**
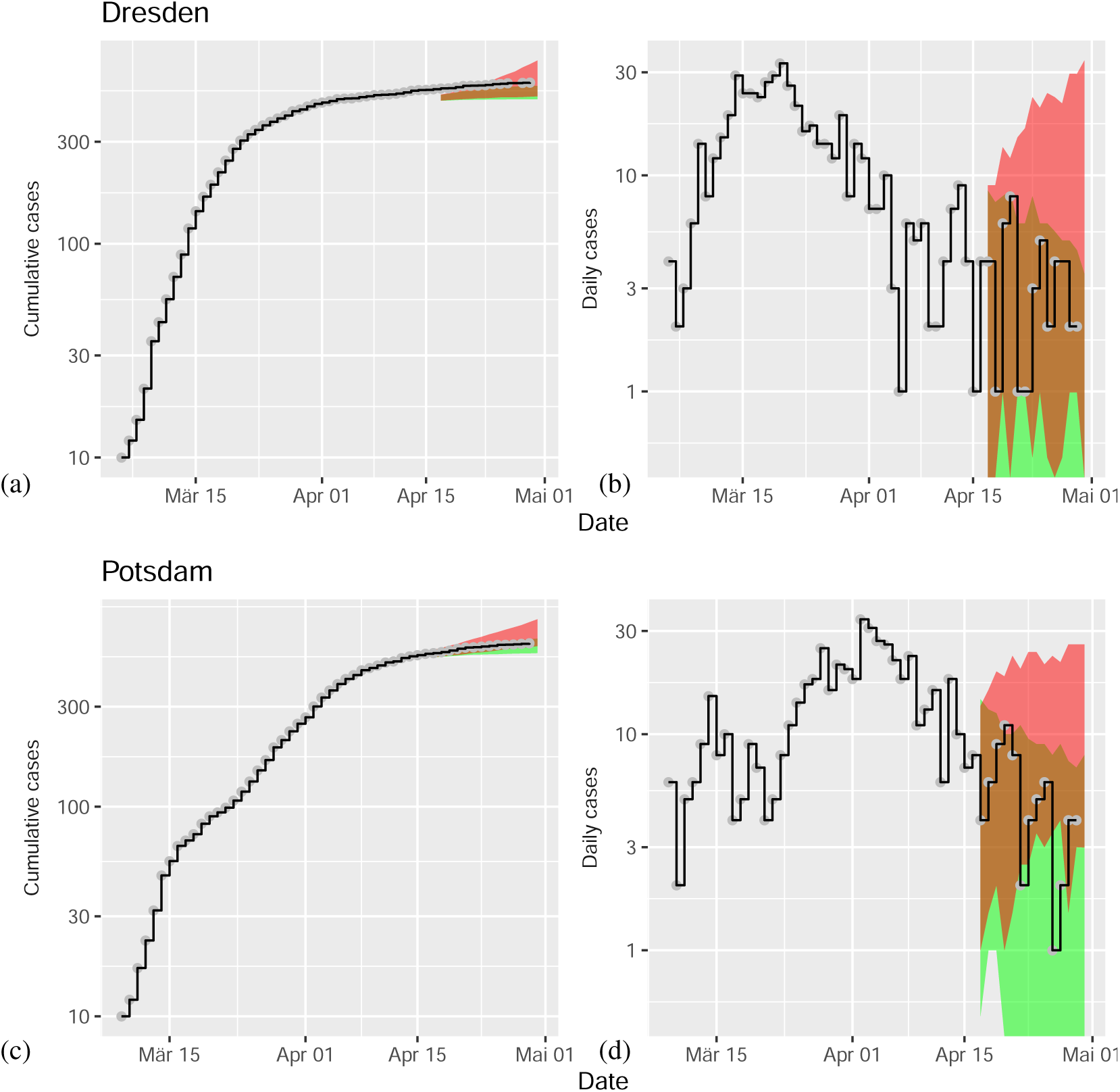
Model predictions for COVID-19 after data assimilation. For details of modeling scenarios I and II see main text and Figure 4.

**Figure S6:**
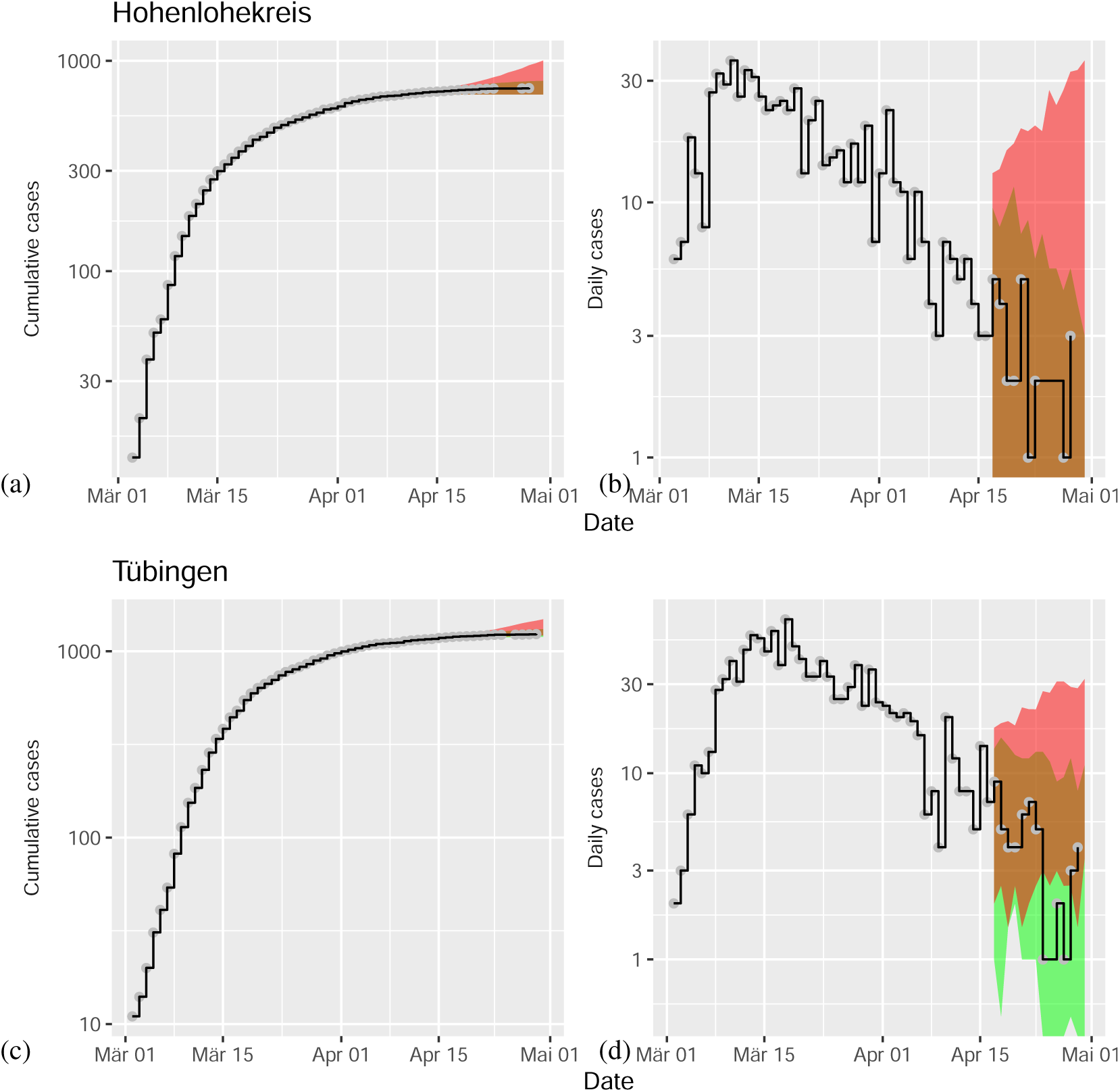
Model predictions for COVID-19 after data assimilation. For details of modeling scenarios I and II see main text and Figure 4.

**Figure S7:**
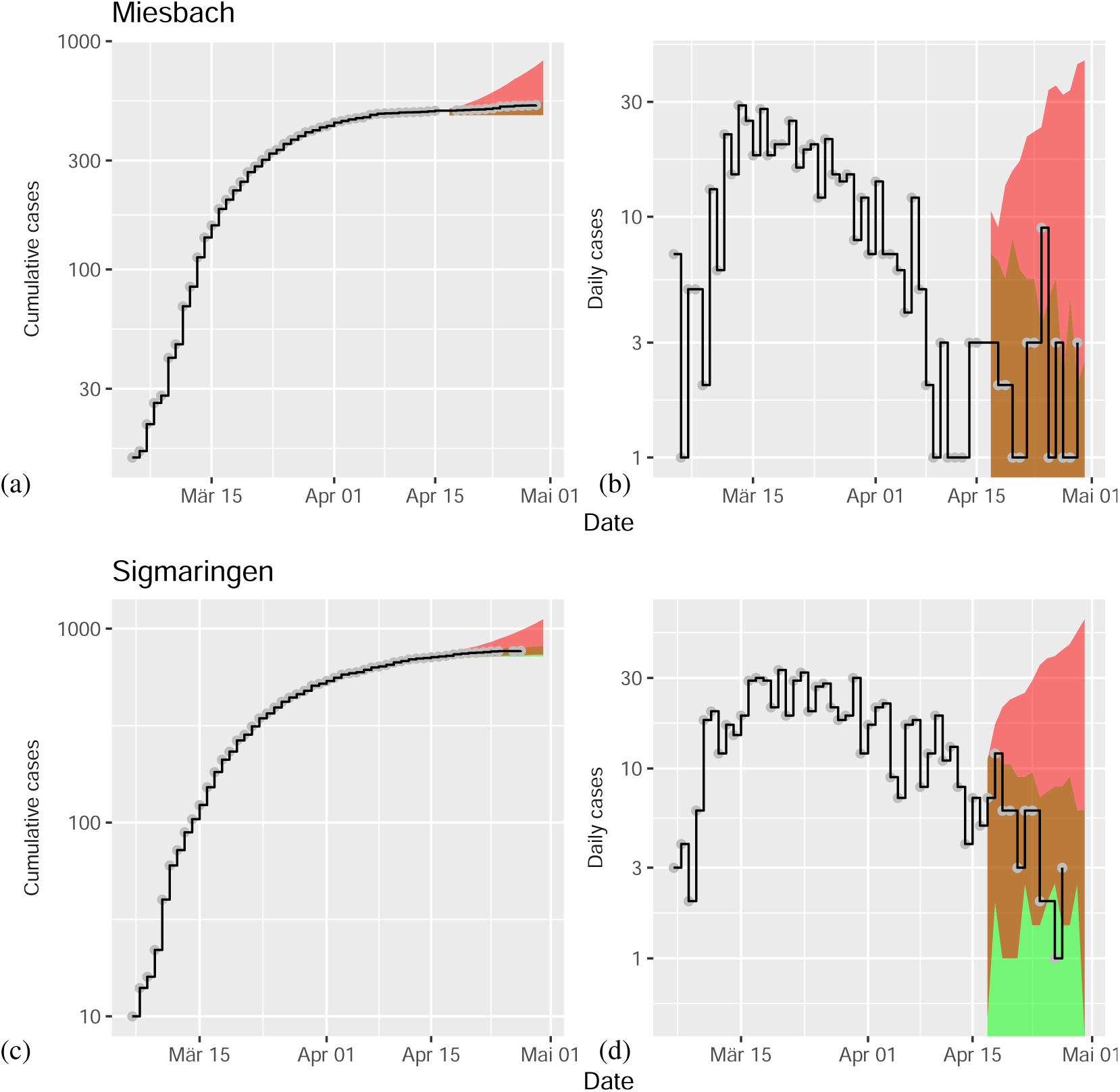
Model predictions for COVID-19 after data assimilation. For details of modeling scenarios I and II see main text and Figure 4.

